# Phenotypic age acceleration through a lens of intersectional inequalities in the German national cohort (NAKO)

**DOI:** 10.1101/2025.02.26.25322953

**Authors:** Enrique Alonso-Perez, Julie Lorraine O’Sullivan, Georg Fuellen, Paul Gellert, Henrik Rudolf

## Abstract

**Background:** Differences in biological aging have been linked to sociodemographic characteristics, but how multiple social inequalities intersect to shape biological aging differences across population subgroups remains unclear. By integrating a perspective of biology of aging with intersectionality theory, we aimed to investigate the joint influence of multiple social determinants on phenotypic age acceleration (i.e., difference between biological and chronological age).

**Methods:** We analysed data from 173,925 participants in the German NAKO study to calculate phenotypic age acceleration. We then created intersectional social strata based on individual sociodemographic characteristics to assess differences in phenotypic age acceleration through an intersectional Multilevel Analysis of Individual Heterogeneity and Discriminatory Accuracy (MAIHDA).

**Results:** All intersectional strata displayed phenotypic age deceleration (i.e., were biologically younger than their chronological age). This advantage was weakest among men without a migration background, living alone and with low socioeconomic status. Substantial discriminatory accuracy of the strata (7.13%) implied intersectional inequalities. Most differences were driven by additive effects, with modest multiplicative effects due to intersectional interactions. We found multiplicative effects representing increased risk for individuals with migration background, not living alone and with medium/high socioeconomic status, or those without migration background, living alone and with medium/low socioeconomic status.

**Conclusion:** Our study provides novel insights on the intersectional stratification of biological aging, highlighting the significance of bio x social interactions for the aging process. Future epidemiological studies should focus on the mechanisms linking multiple social inequalities and accelerated biological aging, using intersectionally-informed targeted interventions that address both social and aging-related inequalities.

**WHAT IS ALREADY KNOWN ON THIS TOPIC:**

- Biological aging varies by sociodemographic factors, with lower socioeconomic status linked to accelerated aging. However, most studies examined single social determinants rather than the interaction effects at their intersections.

**WHAT THIS STUDY ADDS:**

- Using the innovative MAIHDA framework, we identify intersectional disparities in biological aging in a large German cohort.
- While aging differences are largely additive, certain social strata experience amplified disparities due to intersectional effects.

**HOW THIS STUDY MIGHT AFFECT RESEARCH, PRACTICE AND/OR POLICY:**

- Our findings support targeted public health strategies addressing cumulative social disadvantages in aging.
- Future research should integrate intersectional approaches to better understand aging inequalities and design tailored interventions.

## INTRODUCTION

Normal aging is a universal process involving biological, psychological, and social changes that unfold gradually over time.^1^ However, the experience of aging is far from uniform, as individuals may face aging consequences at different rates due to variations in biological processes and social exposures.^2^ ^3^ Chronological age, while a critical risk factor for aging-related morbidity and mortality, fails to capture the heterogeneity of biological aging. Individuals of the same chronological age can exhibit substantial differences in age-related diseases and overall health,^4^ shaped by their social and environmental factors.^5^ ^6^ Adverse exposures, health behaviours, and life-course events can drive biological changes that accelerate aging processes, suggesting the need for a holistic biosocial approach that examines how psycho-social and biological factors interact to influence aging.^7^ ^8^

A growing body of research underscores the profound impact of social determinants on biological aging, with studies consistently revealing stratified associations with sociodemographic factors such as sex/gender, race/ethnicity, and socioeconomic status (SES).^9–12^ For instance, exposure to multiple forms of discrimination has been linked to accelerated biological aging.^13^ Intersectionality theory provides a valuable lens to understand how overlapping social determinants jointly shape health outcomes through structural discrimination and power imbalances.^14^ These cumulative and intersecting (dis)advantages leave biological imprints that intensify over time, influencing aging trajectories in unequal ways.^15^ A recent review emphasized the need for intersectional approaches in aging research, particularly for studying multiply disadvantaged populations where layered inequalities produce unique aging experiences.^16^ Despite its potential to shed light on the biological embodiment of the social environment, intersectionality has been largely overlooked in the study of biological aging and its social stratification.

Biological aging refers to the biological aspects of the deterioration of health and the increase in mortality that occurs with advancing age.^17^ ^18^ Biological age is considered a strong estimator of future health and survival, reflecting the body’s capacity to respond to internal and external stressors. Since it varies significantly among individuals, several models have been developed to estimate biological age. Among these, epigenetic clocks based on DNA methylation (DNAm) emerged as robust predictors;^19^ however, their requirement of large-scale DNA methylation data limits their feasibility for widespread clinical practice. In contrast, serum biomarker-based models such as Biological Age (BioAge)^4^ and Phenotypic Age (PhenoAge)^20^ are a more accessible and efficient alternative. These practical and well-validated aging clocks rely on routinely collected clinical biomarkers, allowing for immediate and cost-effective biological age estimation without the need for additional data collection or complex molecular analyses. By design, these models are strongly associated with risks of aging-related diseases, morbidity, and mortality, making them valuable tools for investigating biological age and its divergence from chronological age—often referred to as biological age acceleration.^21–23^

All biological age measures, whether DNAm-based or serum-based, show a strong socioeconomic and racial/ethnic gradient, indicating accelerated biological aging.^9^ Despite the link between social (dis)advantage and accelerated biological aging, much of the existing research focused on single social determinants, thereby overlooking the interaction effects of intersecting social inequalities. This fragmented approach fails to account for the “double accumulation” of disadvantage experienced both across intersectional social identities and throughout the life-course. Multilevel Analysis of Individual Heterogeneity and Discriminatory Accuracy (MAIHDA) has recently emerged as a cutting-edge method for integrating intersectionality into quantitative health research.^24^ ^25^ By clustering individuals within intersectional social strata based on combinations of their social characteristics, MAIHDA allows to accurately measure and partition the variance in certain health outcomes across strata with better scalability and parsimony than single-level interaction models.^26^ This framework offers a promising approach for applying intersectionality to investigate how cumulative social inequalities are linked to distinct biological aging trajectories. So far, MAIHDA has not been applied to study differences in PhenoAgeAccel.

The present study investigates differences in phenotypic age acceleration across intersectional social strata. Additionally, we aim to identify specific strata where intersectional interactions are associated with accelerated biological aging.

## METHODS

### Data and Sample

The German National Cohort (NAKO) is a prospective population-based cohort study comprising over 205,000 randomly selected participants from 18 study sites across 13 German states.^27^ The study recruited participants aged between 20 and 69 years old with written informed consent and healthy enough for study participation on site. Participants aged >40 years were oversampled, with the baseline data collection taking place between 2014 and 2019. Further details on the study procedures and data quality assessments are published elsewhere.^28^ The study received ethics approval from all centres and was conducted in accordance with the ethical standards from the Declaration of Helsinki. After excluding participants with missing data on strata-defining variables (sex/gender, migration background, education, living alone, or income), and those with insufficient blood biomarkers for imputing PhenoAgeAccel, our final sample included 173,925 participants.

### Outcome variables

Based on Morgan Levine’s algorithm^20^, we calculated PhenoAge with nine biomarkers (albumin, creatinine, glucose, natural log-transformed C-reactive protein (CRP), lymphocyte percent, mean cell volume, red blood cell distribution width, alkaline phosphatase, and white blood cell count) and the chronological age. Since the nine biomarkers were not available for all individuals (missing values ranging from 4.12% for creatinine to 81.22% for lymphocytes), we carried out multiple imputations of missing values. We used 58 out of the 61 available NAKO blood biomarkers as auxiliary variables in the imputation process, since the fully conditional approach demands the maximum available number of lab biomarkers.^29^ Once all biomarkers were imputed, we calculated PhenoAge^20^ with the following formula:

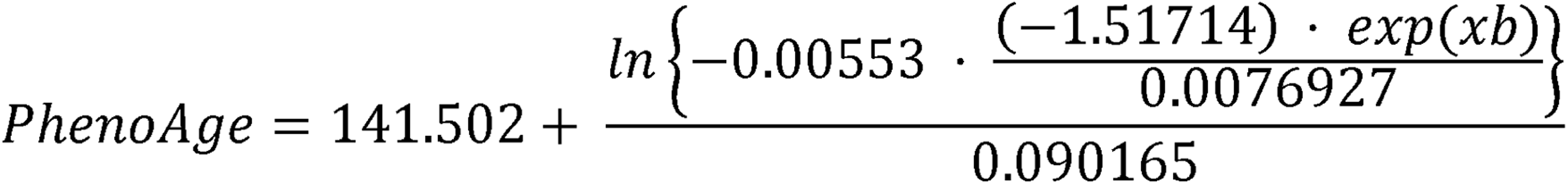

where xb = −19.907 − 0.0336 × Albumin + 0.0095 × Creatinine + 0.1953 × Glucose + 0.0954 × ln (CRP) − 0.0120 × Lymphocyte percent + 0.0268 × Mean cell volume + 0.3306 × Red blood cell distribution width + 0.00188 × Alkaline phosphatase + 0.0554 × White blood cell count + 0.0804 × Chronological age.

Finally, we calculated PhenoAgeAccel as the difference between PhenoAge and chronological age. Positive values imply accelerated aging, whereas negative values reflect decelerated aging.

### Predictor variables

We used four socio-demographic variables to create intersectional strata, based on social determinants that potentially stratify health outcomes.^30^ Sex/gender was categorized as female/male. Migration background was self-reported and classified as yes/no. Living alone was measured via the number of household inhabitants and coded as yes/no. Education level was self-reported and coded as low (primary school), medium (secondary school), or high (university or higher education). Household income was captured by the question “What is the average monthly income in your household?” and was coded as low (<€2,150/month), medium (€2,150–€4,250/month), or high (>€4,250/month). We created 72 unique intersectional strata through all possible combinations of sex/gender (2 categories), migration background (2 categories), living alone (2 categories), education (3 categories) and income (3 categories) (2×2×2×3×3=72).^26^

### Statistical Analysis

We performed a MAIHDA for PhenoAgeAccel with respondents (level 1) nested within intersectional social strata (level 2) based on their individual characteristics.^25^ MAIHDA consists of fitting two sequential multilevel models: first, we fitted an unadjusted null model (Model 1) with a random effect for the intersectional strata, which allows to decompose the variance and calculate the discriminatory accuracy through the Variance Partition Coefficient (VPC).^26^ Analogous to the Intraclass Correlation Coefficient (ICC), the VPC is a measurement of discriminatory accuracy that estimates the between-strata variance in PhenoAgeAccel. Second, we fitted a model adding the stratum-defining variables as main fixed effects (Model 2). We calculated the VPC in Model 2, and, to quantify the between-stratum variance attributable to the additive main effects, we calculated the proportional change in variance (PCV). A PCV < 100% indicates that additive effects of strata-defining variables cannot fully explain the stratum-level variation, implying the presence of multiplicative interactions.^24^ Finally, we analysed the strata-level residuals and their 95% credible intervals (CI) to partition the variance into additive and multiplicative effects, the latter capturing the unique contribution of intersectional interactions. Strata exhibiting two-sided 95% CI that did not include 0 would have statistically significant multiplicative effects, either hazardous (>0, higher PhenoAgeAccel than expected from main effects only) or protective (<0, lower PhenoAgeAccel than expected from main effects only). Further methodological details about MAIHDA are thoroughly explained in a recently published tutorial paper.^26^ We determined statistical significance with a two-tailed p-value < 0.05 for regression coefficients. The statistical software package R (version 4.4.2) was used for statistical analysis.^31^ Descriptive statistics were calculated with the R package “CompareGroups”. All models were fitted using Markov Chain Monte Carlo (MCMC) methods with the R package “brms” as Bayesian regression models (version 2.22). Stratum level estimates are given with 95% credible intervals.

## RESULTS

Table 1 shows the characteristics of the sample, which was balanced with regards to sex/gender. Most respondents did not have a migration background (83.9%) or did not live alone (80.3%). The majority of the sample (44.8%) had a medium income (2,150€ −4,250€), and medium (41.9%) or high (55.5%) education level. Yet, women had lower income levels than men on average. The average blood biomarker levels did not vary substantially by sex/gender. The average PhenoAge was 44.1 years, and the average PhenoAgeAccel was −6.1 years, entailing a younger biological age compared to the chronological age. This phenomenon was even more pronounced among women (−6.9 years compared to −5.4 years for men).

**Table 1.** Descriptive statistics of the study sample, by sex.

| **Table 1** Descriptive statistics of the study sample, by sex |  |  |  |  |  |
| --- | --- | --- | --- | --- | --- |
| **Variable** | **Total**<br>**(N = 173,925)** | **Female**<br>**(N = 86,647)** | **Male**<br>**(N = 87,278)** | **p-value** | **N** |
| Age | 50.2 (12.3) | 50.1 (12.3) | 50.4 (12.3) | <0.001 | 173,925 |
| Migration background |  |  |  | 0.385 | 173,925 |
| Yes | 27,999 (16.1%) | 14,016 (16.2%) | 13,983 (16.0%) |  |  |
| No | 145,926 (83.9%) | 72,631 (83.8%) | 73,295 (84.0%) |  |  |
| Living alone |  |  |  | <0.001 | 173,925 |
| Yes | 34,346 (19.7%) | 18,368 (21.2%) | 15,978 (18.3%) |  |  |
| No | 139,579 (80.3%) | 68,279 (78.8%) | 71,300 (81.7%) |  |  |
| Education |  |  |  | <0.001 | 173,925 |
| Low | 4,521 (2.6%) | 2,773 (3.2%) | 1,748 (2.0%) |  |  |
| Medium | 72,875 (41.9%) | 40,117 (46.3%) | 32,758 (37.6%) |  |  |
| High | 96,529 (55.5%) | 43,757 (50.5%) | 52,772 (60.4%) |  |  |
| Income level |  |  |  | <0.001 | 173,925 |
| <2,150€ | 50,052 (28.8%) | 28,380 (32.8%) | 21,672 (24.8%) |  |  |
| 2,150€ - 4,250€ | 77,968 (44.8%) | 38,794 (44.8%) | 39,174 (44.9%) |  |  |
| >4,250€ | 45,905 (26.4%) | 19,473 (22.5%) | 26,432 (30.3%) |  |  |
| Systolic blood pressure (mm HG) | 127 (10.1) | 122 (9.2) | 131 (9.0) | <0.001 | 173,633 |
| Albumin (g/l) | 42.0 (2.0) | 41.0 (4.1) | 42.0 (1.2) | <0.001 | 98,982 |
| Creatinine (μmol/l) | 72.0 (9.0) | 64.0 (6.3) | 80.0 (8.0) | <0.001 | 166,751 |
| Glucose (mmol/l) | 5.20 (0.6) | 5.10 (0.4) | 5.30 (0.5) | <0.001 | 164,494 |
| C-reactive protein (mg/l) | 1.02 (0.6) | 1.10 (0.7) | 0.96 (0.4) | 0.001 | 104,557 |
| Lymphocytes (%) | 28.8 (4.7) | 29.2 (4.9) | 28.5 (5.2) | 0.001 | 32,667 |
| Mean erythrocyte volume (fl) | 89.7 (2.8) | 90.0 (2.7) | 89.5 (3.1) | 0.001 | 70,680 |
| Erythrocyte distribution (%) | 13.2 (0.8) | 13.2 (0.7) | 13.2 (0.5) | 0.001 | 41,850 |
| Alkaline phosphatase (μkatal/l) | 1.20 (0.2) | 1.10 (0.1) | 1.20 (0.2) | <0.001 | 88,907 |
| Leukocytes (Gpt/l) | 6.12 (1.2) | 6.24 (1.3) | 6.02 (1.1) | <0.001 | 160,982 |
| PhenoAge | 44.1 (9.6) | 43.9 (9.3) | 44.4 (9.7) | <0.001 | 173,925 |
| PhenoAgeAccel | -6.1 (2.6) | -6.9 (2.4) | -5.4 (2.3) | <0.001 | 173,925 |
| Categorical variables are presented as frequencies (percentages), and continuous variables are presented as means with standard deviations (SD) |  |  |  |  |  |
| PhenoAge, phenotypic age; PhenoAgeAccel, phenotypic age acceleration. |  |  |  |  |  |

Figure 1 shows the predicted PhenoAgeAccel for each intersectional stratum from MAIHDA Model 1. One stratum — women with a migration background, living alone, low education and high income — had no observations. While all 71 non-empty strata displayed negative PhenoAgeAccel, substantial between-strata heterogeneity was observed. The lowest deceleration (PhenoAgeAccel = −2.43) occurred in men without a migration background, living alone, with low education and low income, while the highest (PhenoAgeAccel = −7.19) was found in women without a migration background, not living alone, with high education and high income. These findings suggest intersectional inequalities in PhenoAgeAccel across strata (maximum difference = 4.76 years). Table 2 presents results from intersectional MAIHDA models, including the average effects of strata-defining variables and discriminatory accuracy measures (VPC and PCV). The VPC in the null model (Model 1) indicated that 7.13% of the differences in PhenoAgeAccel was explained at the strata level. This represents substantial clustering,^24^ aligned with the heterogeneity in Figure 1. Adding strata-defining variables as fixed effects (Model 2) reduced the VPC to 0.21%. A PCV of 97.29% indicated that most between-strata PhenoAgeAccel differences were due to additive effects, with only 2.71% explained by multiplicative effects (i.e., intersectional interactions).

**Figure 1.**
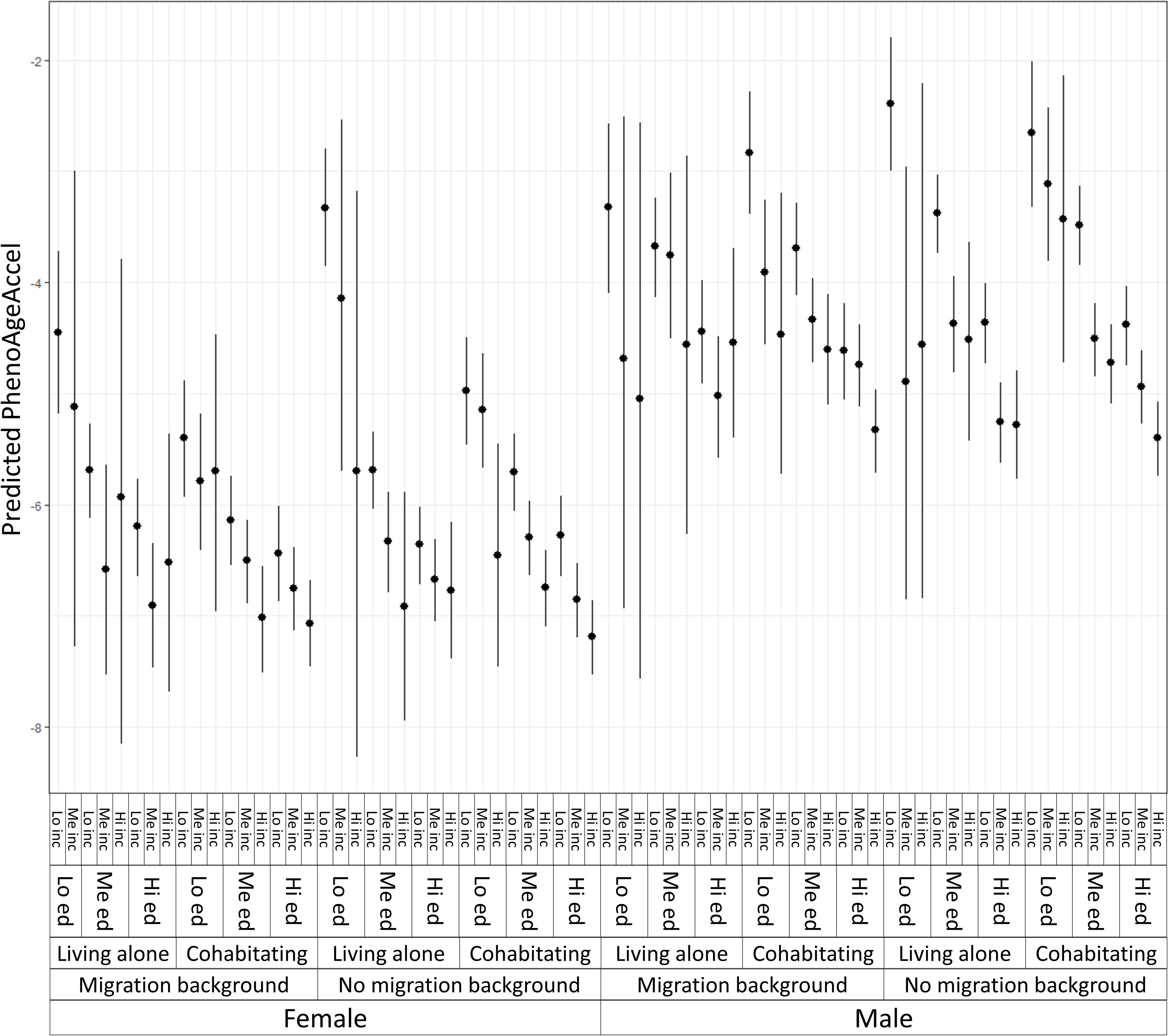
Predicted phenotypic age acceleration (PhenoAgeAccel) for each intersectional social stratum, obtained from MAIHDA model 1.

**Table 2.**
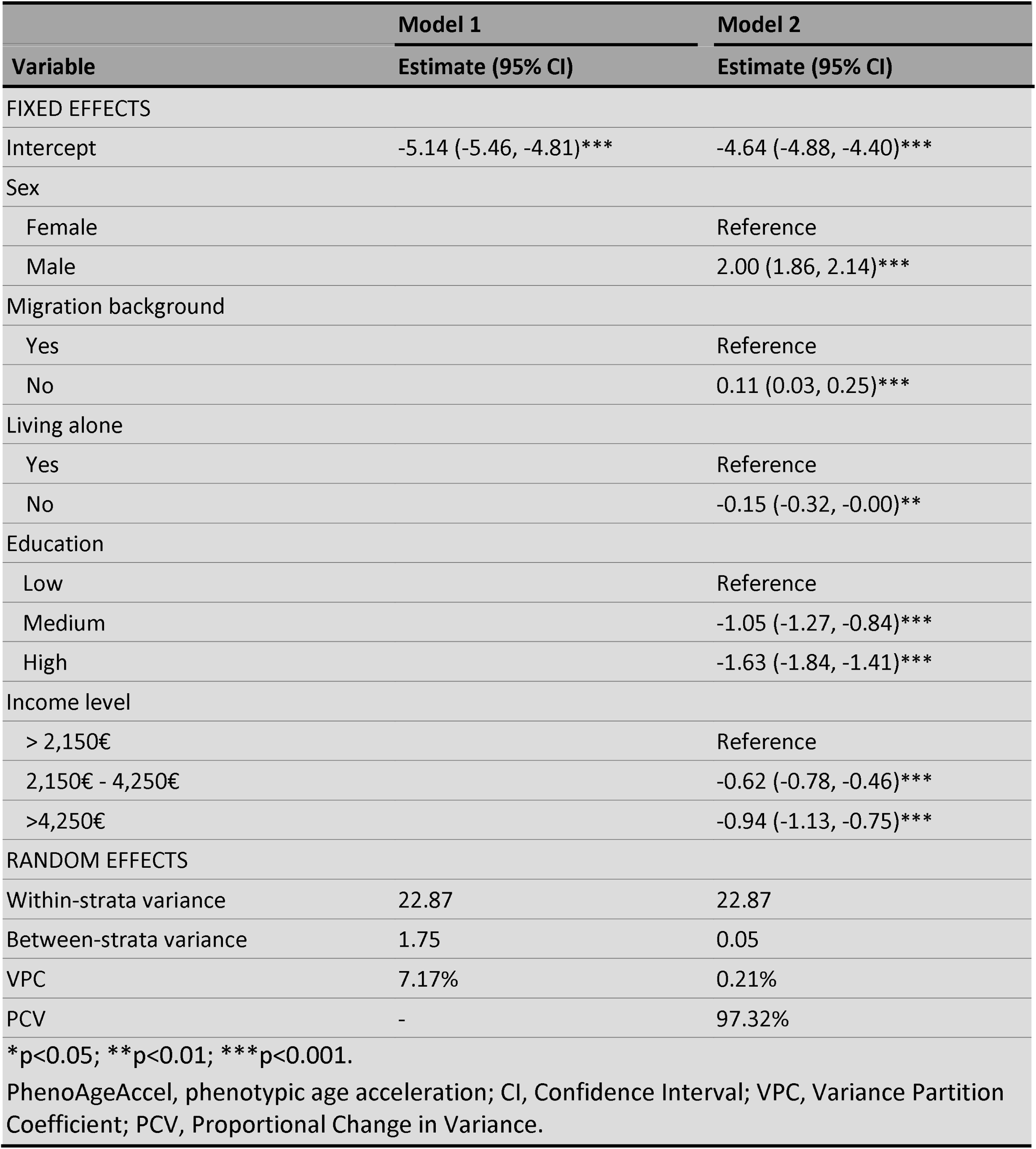
Results from MAIHDA models on PhenoAgeAccel (N = 173,925)

| **Table 2** Results from MAIHDA models on PhenoAgeAccel (N = 173,925) |  |  |
| --- | --- | --- |
|  | **Model 1** | **Model 2** |
| **Variable** | **Estimate (95% CI)** | **Estimate (95% CI)** |
| FIXED EFFECTS |  |  |
| Intercept | -5.14 (-5.46, -4.81)*** | -4.64 (-4.88, -4.40)*** |
| Sex |  |  |
| Female |  | Reference |
| Male |  | 2.00 (1.86, 2.14)*** |
| Migration background |  |  |
| Yes |  | Reference |
| No |  | 0.11 (0.03, 0.25)*** |
| Living alone |  |  |
| Yes |  | Reference |
| No |  | -0.15 (-0.32, -0.00)** |
| Education |  |  |
| Low |  | Reference |
| Medium |  | -1.05 (-1.27, -0.84)*** |
| High |  | -1.63 (-1.84, -1.41)*** |
| Income level |  |  |
| > 2,150€ |  | Reference |
| 2,150€ - 4,250€ |  | -0.62 (-0.78, -0.46)*** |
| >4,250€ |  | -0.94 (-1.13, -0.75)*** |
| RANDOM EFFECTS |  |  |
| Within-strata variance | 22.87 | 22.87 |
| Between-strata variance | 1.75 | 0.05 |
| VPC | 7.17% | 0.21% |
| PCV | - | 97.32% |
| *p<0.05; **p<0.01; ***p<0.001. |  |  |
| PhenoAgeAccel, phenotypic age acceleration; CI, Confidence Interval; VPC, Variance Partition Coefficient; PCV, Proportional Change in Variance. |  |  |

Figure 2 presents the strata-level residuals from MAIHDA Model 2. Most residuals include 0 in their CIs, meaning no statistical significance and suggesting that PhenoAgeAccel between-strata variance was primarily due to additive effects. Only five residuals were significantly different from 0, implying multiplicative effects (Table 3): four strata had hazardous intersectional interactions (positive residuals indicating higher PhenoAgeAccel than expected from the main effects), while one stratum had protective intersectional interactions (negative residual indicating lower than expected PhenoAgeAccel). Hazardous multiplicative effects were observed in strata combining migration background, not living alone and medium/high SES (education and income), or those without a migration background, living alone and with medium/low SES. The only stratum with protective multiplicative effects was comprised by individuals without a migration background, living alone and with medium/high SES.

**Figure 2.**
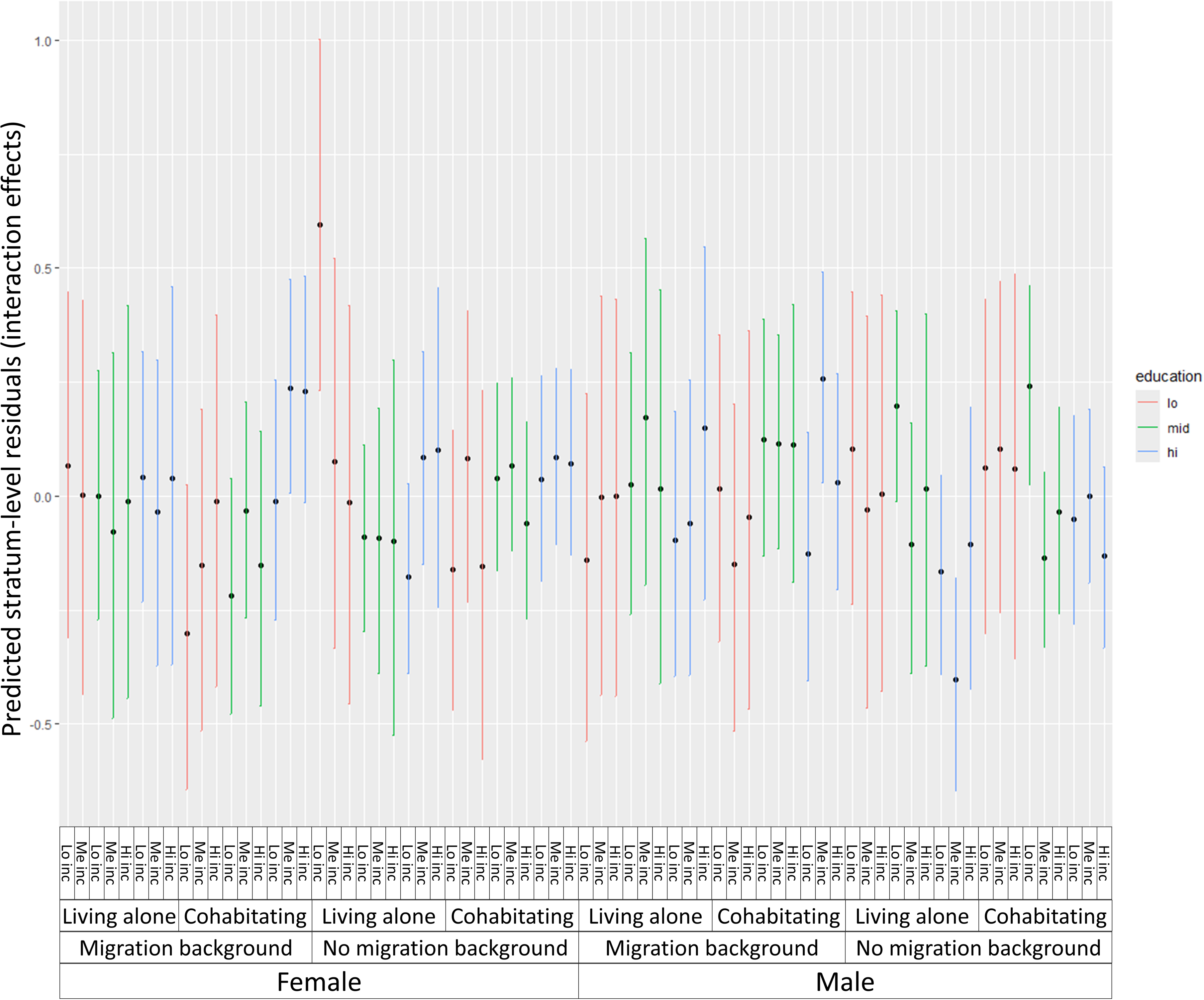
Strata-level residuals for each intersectional stratum, obtained from MAIHDA model 2.

**Table 3.** Composition of the significant strata-level residuals obtained from MAIHDA model 2.

|  |  |  |  |  |  |  |  |  |  |  |  |  |  |
| --- | --- | --- | --- | --- | --- | --- | --- | --- | --- | --- | --- | --- | --- |
| Stratum | Sex |  | Migration background |  | Living alone |  | Education |  |  | Income |  |  | Residual (95% CI) |
|  | Female | Male | Yes | No | Yes | No | Low | Me. | High | Low | Me. | High |  |
| 12111 | X |  |  | X | X |  | X |  |  | X |  |  | 0.60 (0.21, 0.98) |
| 21232 |  | X | X |  |  | X |  |  | X |  | X |  | 0.26 (0.03, 0.49) |
| 22221 |  | X |  | X |  | X |  | X |  | X |  |  | 0.24 (0.03, 0.46) |
| 11232 | X |  | X |  |  | X |  |  | X |  | X |  | 0.24 (0.00, 0.47) |
| 22132 |  | X |  | X |  |  |  |  | X |  | X |  | -0.40 (-0.63, -0.17) |
| The Stratum ID are coded for each digit as follows: Sex: 1 = Female, 2 = Male, Migration background: 1 = Yes, 2 = No, Living alone: 1 = Yes, 2 = No, Education: 1 = Low, 2 = Medium, 3 = High, Income: 1 = Low, 2 = Medium, 3 = High; Me. = Medium education. |  |  |  |  |  |  |  |  |  |  |  |  |  |

## DISCUSSION

In the current study we focus on intersectional inequalities in accelerated biological aging, quantified through PhenoAgeAccel. Using data from 173,925 participants in the German National Cohort (NAKO), we employed the MAIHDA framework to explore intersectional differences in biological aging. By investigating nuanced patterns of social disparity, we found substantial between-strata inequalities in accelerated biological aging, with a maximum difference of 4.76 years in PhenoAgeAccel. Our study offers novel insights into the intersectional stratification of biological aging, highlighting both additive and multiplicative effects of multiple social determinants. The majority of variance in PhenoAgeAccel across strata was due to additive effects, indicating that individual factors like sex/gender, migration background, living alone, education and income independently contributed to differences in biological aging acceleration. However, the presence of multiplicative effects for certain strata suggests that specific combinations of social determinants amplify or mitigate the risk of accelerated aging for these subgroups. The outcomes of our study underscore the critical role of cumulative and intersecting social disadvantages, which, together with biological factors, shape biopsychosocial mechanisms that create differences in the aging process.

Our findings are aligned with previous research linking socioeconomic disadvantage to accelerated biological aging. Studies consistently reported gradients in biological aging associated with income, education and race/ethnicity.^9^ ^20^ ^32^ Particularly, lower income and lower education have been linked to higher age acceleration, consistent with our results showing greater PhenoAgeAccel among individuals with low education, low/medium income and a migration background. However, most studies relied on single-axis frameworks that fail to capture the cumulative and interacting effects of multiple social determinants. By employing MAIHDA with an intersectional lens, we extend this literature and identify strata-specific multiplicative effects, such as the increased risk among individuals with migration backgrounds and medium-to-high SES. These intersectional positions often entail financial pressures, limited access to care resources and adverse health behaviours, which collectively disrupt physiological processes and accelerate the aging process.^33^ ^34^ This underscores how privilege and structural disadvantage intersect to amplify aging disparities, a nuance often missed in traditional approaches.^35^

Accelerated biological aging results from a combination of physiological and psychosocial factors.^36^ Chronic stress, stemming from structural inequalities and discrimination, is a plausible mechanism linking social determinants to accelerated aging.^37^ Its physiological dysregulation consequences (e.g., inflammation, telomere shortening and cellular damage) may explain accelerated aging among disadvantaged intersectional groups.^38^ Our results suggest that stress pathways could be particularly pronounced in individuals with multiple social disadvantages such as low SES and a migration background, who face greater environmental challenges while having fewer psychosocial resources. This can lead to chronic stress, reduced coping ability, prolonged psychobiological activation, and a reduced recovery capacity.^39^ Conversely, protective effects in certain high-SES strata may reflect resilience mechanisms related to more privileged social positions, including access to healthcare, healthier lifestyles, and stronger social support, which may buffer the impact of stress,^40^ even in case of migration background— though this finding is sex-specific. Most importantly, these physiological and psychosocial factors intensify when social (dis)advantages overlap, underscoring the need of an intersectional perspective. Examining single social determinants in isolation risks overlooking the joint effects that emerge at their intersections.

The social hallmarks of aging, such as adverse life events, low SES, minority status and adverse psychological status, play a significant role in the aging process.^2^ These social determinants are deeply intertwined with individuals’ socioeconomic positions, social networks, and access to resources, which can either accelerate or protect against biological aging. From an intersectional perspective, it is crucial to consider how these social hallmarks interact with multiple dimensions of disadvantage to produce unique experiences of aging across different social groups. Our study is the first to incorporate an intersectional approach to the social hallmarks of aging, yet future research should explore how the intersection of all the social factors proposed by Crimmins affect age-related outcomes. Aging is not a uniform experience, but one shaped by the overlapping effects of social and biological factors, hence the integration of biopsychosocial and intersectional approaches into studies of aging is crucial. By doing so, we can better understand the full scope of aging disparities and inform more effective, targeted interventions aimed at mitigating the negative impacts of social disadvantages.

Our study has several strengths that include the use of a large, nationally representative cohort, the extensive availability of serum biomarkers for a large sample, and the application of MAIHDA to rigorously assess intersectional effects. However, our findings should be interpreted in light of several limitations. First, the cross-sectional design of the study and the correlational nature of MAIHDA limit causal inferences. Longitudinal and interventional studies are needed to establish temporal relationships between multiple social determinants and biological aging, with a particular focus on testing the biopsychosocial mechanisms that create such effects. Second, while PhenoAge is a validated measure of biological aging, it does not fully capture all dimensions of the (clinical) biological aging processes, such as cognitive decline or frailty. Future research analysing the social determinants of biological aging should incorporate measures of functioning limitations, to achieve a more comprehensive view on the aging process and its social stratification. Finally, the reliance on self-reported data for certain variables, such as income and education, may introduce reporting bias. Nonetheless, the NAKO study was rigorously planned and carried out with a randomly selected, multi-centre cohort, hence it is plausible to assume that the overall reporting bias is minimized.

The identification of intersectional disparities in biological aging has critical implications for public health and policy, underscoring the importance of addressing social inequalities in health policy and intervention design. Our findings suggest that targeted interventions should prioritize stress reduction and resource allocation in multiply disadvantaged populations to mitigate accelerated aging. For example, policies promoting equitable access to healthcare, income support programs, and anti-discrimination measures may mitigate the accelerated aging observed in high-risk strata. Additionally, public health initiatives could incorporate culturally tailored stress management and mental health programs to address the unique challenges faced by migrant populations.

From a methodological perspective, our study highlights the importance of integrating intersectionality into quantitative longevity research. The use of MAIHDA allowed us to quantify both additive and multiplicative effects of social determinants, providing a more nuanced understanding of health inequalities. Future research should expand intersectional analyses to include other social determinants that may be linked to accelerated biological aging, such as occupational class, ethnicity or neighbourhood deprivation. Additionally, policymakers and researchers should consider employing similar frameworks to address disparities in other health and aging outcomes, such as chronic disease incidence and mortality.

## CONCLUSION

Our study demonstrates the intersectional stratification of biological aging, emphasizing the need for a biopsychosocial approach to address aging-related health disparities. These findings highlight the urgency of addressing social inequalities to mitigate accelerated aging and promote health equity across diverse populations. We demonstrate the utility of applying an intersectionality-informed framework in public health research, which helps to identify subgroups at higher risk for the development of tailored population-level measures.

## Acknowledgements

We thank all participants who took part in the NAKO study and the staff of this research initiative. We would also like to thank all members of the Einstein Center Population Diversity (ECPD). We acknowledge the Projekt DEAL for enabling and organizing Open Access funding.

## Contributors

EAP, PG and GF conceived the study. EAP and PG carried out the data application process. HR performed the statistical analysis, and EAP drafted the manuscript. EAP, JLOS, GF and PG contributed to results interpretation. All authors critically revised the manuscript and approved the final version.

## Patient consent for publication

Not required.

## Ethics approval

The NAKO was approved by the ethical review committees of all participating NAKO study centres. Informed consent was obtained from all participants.

## Provenance and peer review

Not commissioned; externally peer reviewed.

## Data availability statement

The dataset used in the current study is not publicly available due to privacy concerns, but they can be obtained for free upon request via the NAKO transfer hub: https://transfer.nako.de/transfer/index.

